# Delay in Clearance of Labeled Protons Post-Acute Head Trauma Utilizing 3D ASL MRI (Arterial Spin Labeling) a Pilot Study

**DOI:** 10.1101/2023.10.11.23296876

**Authors:** Charles R Joseph, Jubin Kang, Bryce N Grohol, Marija Zivcevska, Joshua Lenke, Ethan Dean Rich, Connor James Arrasmith, Ian Shepherd Dorman, Bradley Waman Clark, Kim Love, Ben Ferry, Mark E Rolfs

**Affiliations:** Liberty University College of Osteopathic Medicine

## Abstract

**Background:** The study aims were to correlate acute head injury cognitive changes with ASL-MRI reduced glymphatic clearance rate (GCRs) and determine GC improvement with recovery. Concussive-brain injury disrupts the blood brain barrier (BBB) and reduces cMTT (capillary mean transit time) and GCRs. Concussion is clinically diagnosed utilizing history and exam findings. ASL-MRI assesses brain perfusion ingress and outflow.

**Methods:** 3D TGSE (turbo-gradient and spin echo) PASL (pulsed arterial spin labeling) 3T MRI with 7 long TI’s (time to inversion) assessed the GCRs (slope of the linear decay of signal vs. time) of labeled protons 2800-4000 ms post-labeling in bifrontal, bitemporal, and biparietal regions within 7 days of mild acute traumatic brain injury and after clinically cleared to return to usual activities. The Sport Concussion Assessment Tool Version 5 (SKAT5) and Brief Oculomotor/Vestibular Assessment (administered by sports physicians) evaluated injured student athletes’ cognitive function prior to ASL MRIs.

**Results:** Pilot study demonstrated significant GCRs improvement (95% [CI] -0.06 to -0.03 acute phase; to [CI] - recovery [CI] 0.0772 to -0.0497 ; P <0.001 in Frontal lobes; and Parietal lobes (95% [CI] -0.0584 to -0.0251 acute; [CI] -0.0727 to - 0.0392 recovery; P = 0.024) in 9 head injured athletes (8 female 1 male mean age 20). 6 age/activity matched normal controls (4 female 2 male mean age 22) were also compared.

**Conclusion:** Acute head trauma disrupts the BBB reducing GCR measured using this 3D ASL MRI technique. ASL MRI is a potential noninvasive biomarker of acute brain injury and subsequent recovery.

**Key Message:** Objective measure of post mild TBI recovery has remained elusive as conventional anatomic imaging techniques and biomarkers are not sensitive. This pilot study demonstrates the potential of leveraging alterations in brain perfusion in the late phase capturing both delayed capillary perfusion and retained free fluid clearance from the brain, both the result of blood brain barrier leak from the acute trauma. Our noninvasive ASL MRI technique identified both anatomic site-specific delay in clearance acutely as well as restoration of normal flow post recovery. This time and cost-efficient noninvasive technique may, with additional validation, provide a needed objective measure for identifying physiologic changes post-acute injury and upon clinical recovery.

## Introduction

Traumatic brain injury (TBI) is a major worldwide health concern affecting an estimated 369 per 100,000. ^1-3^. 90% of injuries are mild (mTBI) defined as Glasgow Coma Scale of 13-15/15. Sports related injuries account for an estimated 17% of injuries, however the number of cases is underestimated as the epidemiologic data is derived from hospital ^4^ admissions excluding those managed conservatively. All cause and severity indirect and direct TBI costs were estimated to be $93 billion in 2019 in the US alone, not including lost wages and unemployment. The latter predominantly affects those with moderate or severe TBI ^3^. The sequela after even mild injury may include overt or subtle emotional, cognitive, and endocrinologic dysfunction ^5,6^.

Diagnosis of sports related acute mild TBI (mTBI) is reliant on cognitive and physical exam inventories such as Sport Concussion Assessment Tool, 5th edition (SKAT5) obtained at the time of injury, and Vestibular Ocular Motor Screening (VOMS) ^7,8^. If these tests are positive for a concussive injury, athletes are held out of related activity until symptoms abate and the above tests revert to baseline. Routine CT brain and anatomic MRI imaging studies are typically normal ruling out structural injury in the vast majority of patients with mTBI but are of little value in addressing the altered physiology due to damaged blood brain barrier (BBB) ^9-10^. Although sensitive to BBB fluid dynamics, MRI Diffusion Tensor Imaging (DTI) is limited by long scan times of 1-2 hours, and dynamic contrast enhancement (DCE) employs gadolinium contrast. Given the potential for contrast deposition within the nervous system with unknown long-term consequences, the technique is not suitable for multiple repeat studies. Functional MRI (fMRI) is also limited by cost and long scan times, which make routine usage impractical ^11-15^.

Our approach is to leverage physiologic changes resulting from blood brain barrier (BBB) leak inherent to acute TBI and repair thereof upon recovery^16^. The BBB leak is a direct result of the blunt force injury and subsequent inflammatory changes that may develop ^16-21^. The result is local alteration in the normal vascular perfusion affecting the capillary mean transit times (cMTT) which is the measure of time blood flows through the capillaries in a given brain region^22^.

The concept of glymphatic clearance rate (GCRs) measures the time dependent slope of the reduction in labeled proton signal in the late phase of perfusion^23^. The constituent labeled proton signal in the late perfusion phase is from both labeled blood inflowing during the capillary mean transit time (cMTT) and presence of residual labeled free fluid present in the interstitium by diffusion. Reduced GCRs (post BBB injury) is caused by delayed cMTT and increased labeled free fluid trapped in the interstitium within injured brain regions. The contribution of each tissue component (white matter, gray matter, blood, and free fluid) within the region of interest at specific post labeling delay times (PLD) of the total signal average is dependent on their respective T1 values [figure 1]. Solving the T1 Bloch equation M_t_ = M_max_(1-e^-t/T1^) for each brain component using their respective T1 values at 3T, and perfusion component data from Li et al clearly shows the component signal in the late phase of perfusion is residual capillary flow plus residual labeled fluid in the region of interest^24^. The choice of timing of the 7 PLDs was based on this combined data which minimized all but the 2 components desired: residual free water and labeled capillary fluid in the interstitium. Note the signal from white matter (<1% at 3X the T1), gray matter (14% at 2X its T1) are minimized by ASL background suppression/subtraction. Thus, the remaining residual signal in the long PLD’s is from residual free labeled water and to capillary labeled blood. Blood brain barrier leak results in delayed cMTT and disruption of glymphatic flow, both demonstrable in the latter portion of the perfusion cycle by ASL MRI. The persistence of labeled protons within the ROI can be graphed versus time, the slope representing GCRs is normally negative (fluid flow out) but is flat or slightly positive in BBB injury (retained fluid) ^10,16,17,22^. White matter fluid flow is delayed regionally by BBB leak to a greater extent than in associated gray matter ^12,25^. 3D ASL MRI is a noninvasive and available sequence on most 3T and above MRI scanners. By labeling protons in the neck, endogenous contrast is produced whose signal can be measured at selected delay times (TI) post labeling (PLD) during the perfusion cycle ^25-29^. By choosing long delay times at 2800-4000 ms, only the residual labeled protons from the delayed capillary phase of perfusion and free fluid are measured in the region of interest (ROI) [fig 1]. The ROI, although hand drawn, encompasses a large region whose volume and shape are held constant, with the same slice chosen for each of the 7 determinations. By measuring signal averages sequentially in ^23^ chosen white matter regions late in the perfusion cycle, the slope of the linear analysis reflects the GCRs ; we refer to^22, 29-33^ this as the GCR slope. Blood brain barrier leak results in a reduced GCRs compared to normal. The GCRs reduction is not specific for head injury but occurs in any disease process damaging the BBB such as Alzheimer disease. By using this approach in a pilot study, we found reduced labeled proton clearance rates in subjects with Alzheimer disease ^27^ (where BBB leak develops early in the process) compared to normal age matched control subjects. Given the presence of BBB leak in mTBI, the possibility of extrapolating the ASL MRI technique was explored. The short scan time of 20 minutes, low cost, and relative safety (no gadolinium contrast) make this a potentially attractive diagnostic biomarker to investigate.

**Figure 1.**
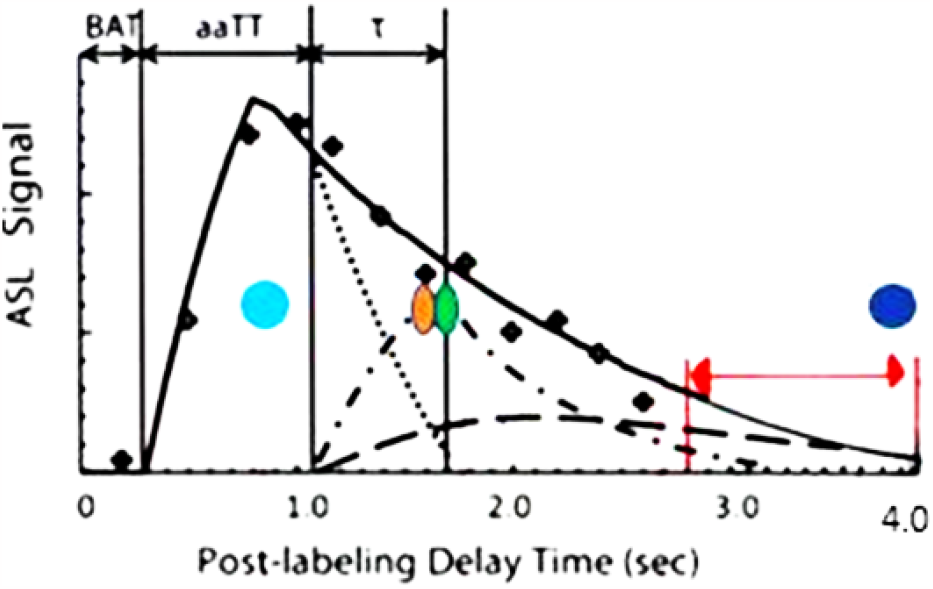
Components of Cerebral Perfusion versus Time. Composition of the ASL signal/time into its various components: arterial, capillary, and extra capillary spaces. TI (acquisition) times utilized in this study are between the red arrow limits. Note the signal composition is capillary and extra-capillary (interstitial) water, the latter is normally cleared by the glymphatic system 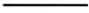. total composite signal vs time 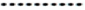 arterial signal contribution vs time intracapillary signal contribution vs time Extracapillary fluid signal contribution vs time. 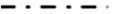 intracapillary signal contribution vs time 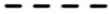 Extracapillary fluid signal contribution vs time. T1 times (63% signal decay) of major signal contributors indicated by the colored dots. Magenta dot = 800-850 msec T1 of white matter. Orange dot = 1650 msec T1 of blood. Green dot = 1700msec T1 of gray matter (all values are for 3T) Dk blue dot = 3800msec T1 of water (CSF fluid); (all values are for 3T) BAT = bolus arrival time; aaTT =artery-artery arrival time; τ = peak capillary arrival time Adapted with publisher permission (Wiley) from reference 24.

## Methods

### Study subject and control recruitment

In this case-controlled study, potential subjects between 18-30 years of age were recruited following an acute mTBI. Nine major university student athletes (1 male, 8 female, ages 19-22) with mTBI and no other significant underlying disease, were studied within 1 week of injury, and then again upon clinical recovery. The study commenced in late 2021 and concluded in the spring of 2023. All subjects gave written consent to participate. One subject had 2 head injuries within 3 weeks and was studied twice before the clinical recovery scan. A control group of age and activity matched volunteers (2 males, 4 females, ages 19-23) was also obtained for comparison (Table 1). Potential variables include location of injury, severity of impact, duration of altered consciousness or confusion, and evidence on FLAIR sequence of new or old structural injury. Potential confounders include additional underlying disease such as chronic CNS disease or injury, systemic inflammatory or chronic illnesses. Potential biases include young athletic heavy subject pool, and inclusion of mild acute brain injury only subjects. The former bias was justified by the otherwise healthy cohort reducing illness confounders and the latter justified by efforts to determine test sensitivity. The study size was extrapolated from our initial pilot study using ASL MRI which demonstrated statistically significant differences (power of 0.8) in 3 Alzheimer disease subjects compared to age matched controls^23^2. Given the inciting presence of BBB leak in both groups (AD and TBI), the numbers seemed justified for an exploratory pilot study. The control group size was also extrapolated from our previous AD pilot study control group clearance rate results^23^.

**Table 1.** Subject Demographics and Acute Injury SKAT5 and VOMS Scores.

| Cohort (n)<br>sex %<br>distribution<br>mean age<br>(MA)<br>(range) | D # | Sport | Date of<br>HI | SCAT5<br>Symptom<br>Quantity | SCAT5<br>Symptom<br>Severity | VOMS<br>$\Delta^*$ | Brain<br>region<br>affected | Days<br>between<br>injury<br>and<br>recovery<br>scans |
| --- | --- | --- | --- | --- | --- | --- | --- | --- |
| mTBI (9)<br>M:F = 1:8<br>11.1% : 88.9%<br>MA=20.2 yrs.<br>(1.2) | 1 | S/DIVING | Nov-21 | 9 | 23 | >3** | BP, LF | 56 |
|  | 2 | Cheer lead | Jan-22 | 10 | 22 | >3** | BF | 27 |
|  | 3 | VB (HI X 2) | Jan-22 | 10 | 22 | >3** | BF, BP | 28 |
|  |  |  | Feb-22 | 10 | 23 | >3** | BF, BP |  |
|  | 4 | WLAX | Feb-22 | 7 | 10 | 6 | BF | 28 |
|  | 5 | WLAX | Sep-22 | 14 | 42 | >3** | BT, BP | 21 |
|  | 6 | MSOC | Sep-22 | 12 | 32 | >3** | BF | 48 |
|  | 7 | WLAX | Feb-22 | 16 | 34 | >3** | BF | 21 |
|  | 8 | FH | Oct-22 | 15 | 36 | 5 | B(T,F,P) | 17 |
|  | 9 | VB | Oct-22 | 10 | 20 | 6 | LF | 28 |
| Control (6)<br>M:F = 2:4<br>33.3%:66.7%<br>MA=21.8 yrs.<br>(2.1) | 1 | S/diving | None |  |  |  |  |  |
|  | 2 | SB | None |  |  |  |  |  |
|  | 3 | Basketball | None |  |  |  |  |  |
|  | 4 | VB | None |  |  |  |  |  |
|  | 5 | runner | None |  |  |  |  |  |
|  | 6 | Track sprint | None |  |  |  |  |  |

**S/diving** =swim/diving, **VB**= volleyball, **WLAX** = women's lacrosse, **MSOC** = men's soccer, **FH** =field hockey, **SKAT 5** =sport concussion assessment tool 5th edition, **VOMS** = vestibular oculomotor screening. B = bilateral F= frontal, P = Parietal, T = temporal, L= left \*The higher post injury score compared with baseline indicates high likelihood of a cerebral concussion. \*\*VOMS score $\geq 3$ (scale of 1-10 assesses headache, dizziness, nausea, fogginess) supports concussion diagnosis. Sensitivity = 64% specificity = 74%

### Subject eligibility

Following an athletic injury that could precipitate a mTBI, an athletic trainer performed an initial evaluation using the SCAT 5. If these results were suggestive of a concussion, the athlete was further evaluated by a sports medicine physician within 48 hours for a definitive diagnosis ^5, 34-36^. The physician reviewed the SCAT5, obtained a history, and performed a physical exam. The physical exam included a cranial nerve exam, and if warranted, a Vestibular Oculomotor Screening (VOMS) as well (Table 1). In addition, osteopathic screening and manipulative treatments were performed in some cases. Per convention, the injured athletes were removed from sport participation and instructed to employ as much cognitive rest as possible, including academic accommodations, limitations to screen-time, et cetera, along with strict physical rest. The athletic training staff followed their recovery progress and would begin a standard five-day return-to-play (RTP) protocol under physician supervision once the athlete was asymptomatic. After the athlete had successfully returned to their academic work and the RTP protocol was completed without symptom recurrence, they were medically cleared to resume full participation in their sport. No financial or other incentives were offered for participation. To be included in the control group, age matched, and as closely as possible, athletic activity or exercise matched uninjured volunteer subjects. Four of the six volunteers were division one athletes, and two of the six students were former athletes and currently physically active.

### DEI

All willing and consented student athletes with acute mild TBI participated in the study, including 8 (89%) females, 1 (11%) male, athletes aged 19-24 years at the time of injury regardless of socioeconomic, race or insurance status, or sport from November 2021 to December 2022. Normal activity matched subjects were recruited 2 (33% male, 4 (66%). Three women of varying career stages are included in the authorship team.

### Patient and public involvement

Given the nature of the study requiring expertise in MRI and Sports medicine as it relates to acute TBI clinical assessment, patient input was essential in determining their subjective clinical recovery, and thus timing of recovery MRI scans. Public representatives on the CENTRA IRB committee made protocol suggestions during the approval process.

### Exclusions

Subjects without witnessed head injury, those with non-removable metal piercings or MRI incompatible implants, claustrophobia, heart disease, or stroke history were excluded from the study.

### Imaging and data analysis methods

Using the 3D TGSE PASL MRI protocol previously published, seven ASL sequences using TI’s (time to inversion) starting at 2800 ms and at 200 ms intervals to 4000 ms were obtained ^23^. In addition, a FLAIR axial sequence was obtained. Total scan time was 18½ minutes. The 3D images were then formatted into 4 mm contiguous axial slices from which a single slice at the same level of all ASL sequences were chosen. Post-processing was completed using perfusion images which were transferred to PACS then using the ROI tool, a slice specific constant volume and location for each brain region were studied (4mm axial slices). Homologous temporal lobe ROI was 650 mm^2^ individually obtained just above the temporal horn for all subjects. Bilateral frontal lobe ROI was 1150 mm^2^ individually just above the lateral ventricles. Bilateral parietal lobes ROI was 760 mm^2^ individually at the same level as the frontal lobe ROI but posterior (Figure 2). By choosing a large ROI in each brain region, avoiding subarachnoid and ventricular spaces, reduced potential sources of error related to contamination with extra-parenchymal fluid and signal voxel-to-voxel signal variability. Signal averages were transferred to a spreadsheet and graphed vs time (Figure 3). Since the data collection occurs in the late phase of the perfusion cycle (egress of blood and fluid), close approximation by linear analysis can be applied with the slope of ^14,23^ the line corresponding to the clearance of labeled protons. The acute injury and recovery results were then pooled and compared statistically. All subjects were scanned in the midafternoon to early evening hours and had withheld alcohol and caffeine for the preceding three hours before their study.

**Figure 2.**
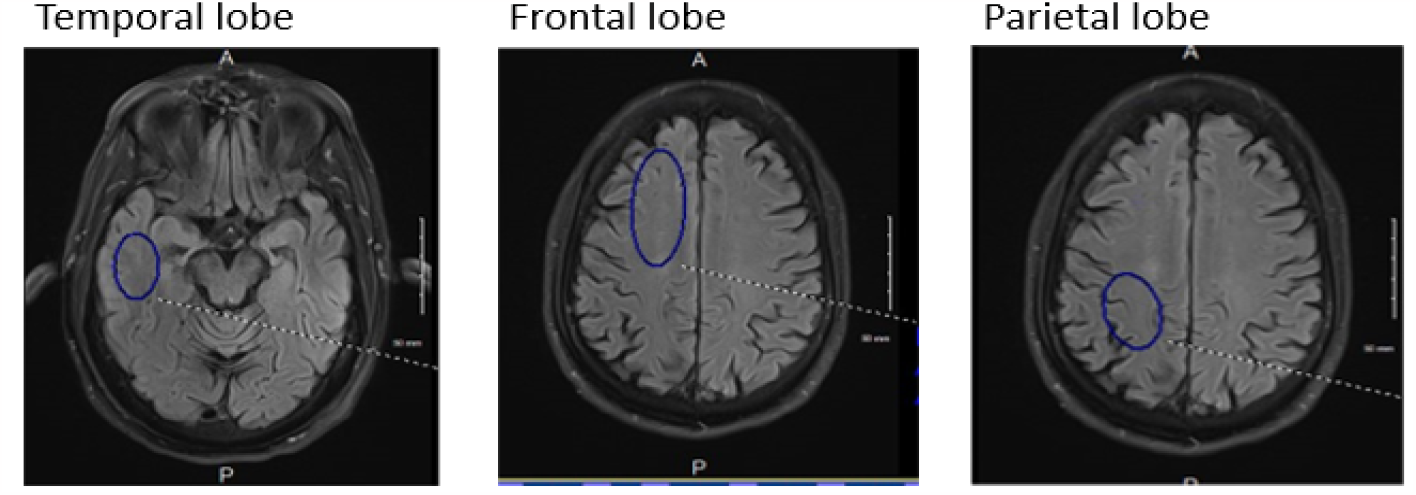
Selected brain regions for ROI signal average. The blue oval contains the region of signal average: temporal lobe 650mm^2^; frontal lobe 1150mm^2^; and parietal lobe 760mm^2^. Homologous contralateral determinations were also obtained.

**Figure 3.**
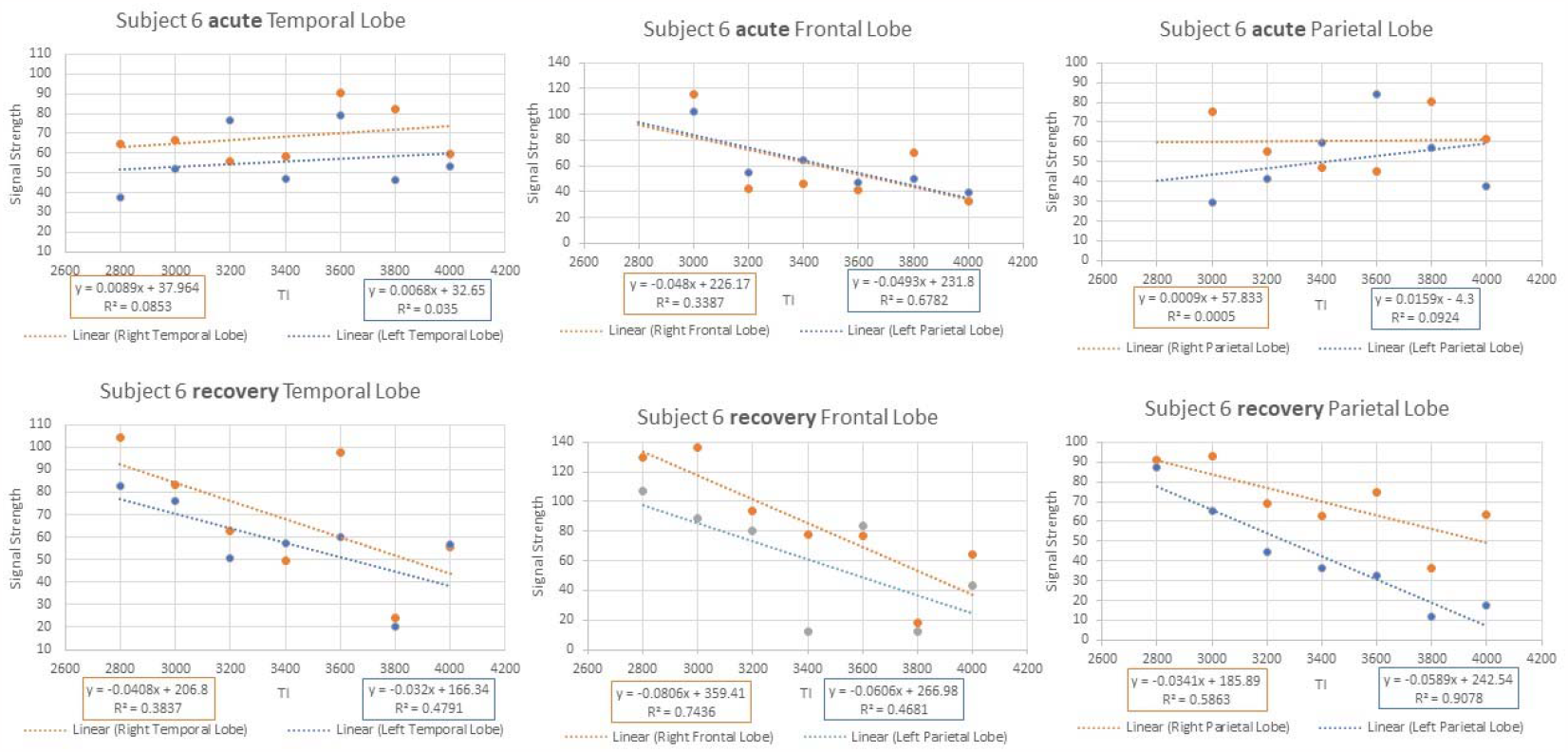
Example of signal averages vs. time. Acute phase temporal lobe and right Parietal lobe show GCR of zero or positive. The same regions upon recovery show negative GCR slopes within the normal range.

### Statistical methods

Gender and age were summarized in each group using frequency distributions and summary statistics. ROI were analyzed in separate models. Because each subject had nested bilateral measures of GCR slope which were repeated under injury and recovery conditions, in order to compare the GCR slopes for the two conditions within mTBI subjects, a linear mixed effects model with random intercepts for subject and lateral side was used initially. After finding minimal variability between the two lateral measures within each subject for all three ROIs, this random intercept was removed from the model (leaving only a random intercept for subjects). Similarly, when comparing mTBI subjects to control subjects, a linear mixed effects model was used with an intercept for each subject in order to account for bilateral nesting of GCR slopes within subjects. Visualization of model residuals indicated approximate normality in all cases. IBM SPSS v 29 was used for all statistical analyses.

## Results

See appendix for the full clearance rate results for all subjects (acute injury and recovery as well as controls).

The results from the three linear mixed effects models comparing the acute and recovery slopes of the line correspond to the clearance of labeled protons over time (Table 2) This table includes estimated marginal means, differences, standard errors, and statistical test results. that the measures are different.

**Table 2.** Results of Linear Mixed Effects Models Comparing Acute and Recovery Labeled Residual Proton Slopes. Table 2 shows that there are statistically significant differences at the 0.05 level of significance with associated confidence intervals for both frontal and parietal lobes (pooled results); in both cases, the acute slopes are positive or less negative than the recovery slopes. The pooled temporal lobe results acute and recovery showed no evidence that the measures are different.

| Lobe | Time | Mean | SE(M) | Difference | 95% CI (Diff) |  | t | df | p |
| --- | --- | --- | --- | --- | --- | --- | --- | --- | --- |
| Temporal | Acute | -0.0545 | 0.0082 | 0.0052 | -0.0115 | 0.0220 | 0.64 | 28.8 | 0.526 |
|  | Recovery | -0.0598 | 0.0084 |  |  |  |  |  |  |
| Frontal | Acute | -0.0462 | 0.0062 | 0.0172 | 0.0083 | 0.0261 | 3.96 | 28.1 | < 0.001 |
|  | Recovery | -0.0635 | 0.0062 |  |  |  |  |  |  |
| Parietal | Acute | -0.0417 | 0.0075 | 0.0142 | 0.0021 | 0.0264 | 2.39 | 28.1 | 0.024 |
|  | Recovery | -0.0559 | 0.0076 |  |  |  |  |  |  |

Figure 4 includes means and standard error bars of the slopes for each of the three lobes at both timepoints.

**Figure 4.**
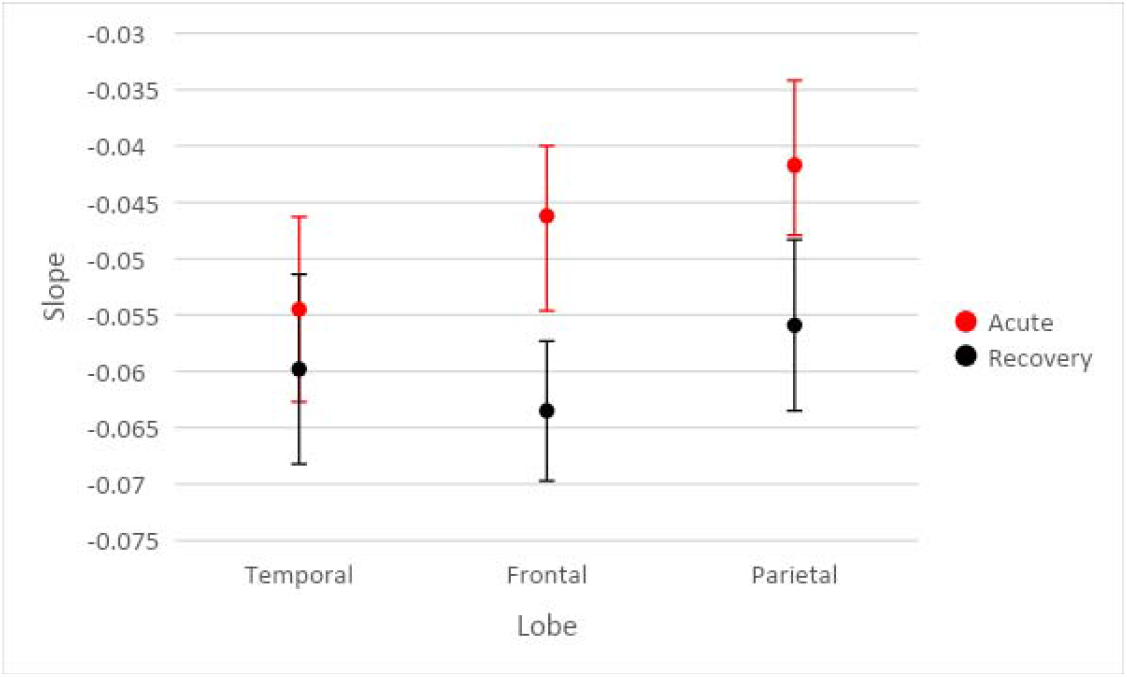
includes means and standard error bars of the GCR for each of the three lobes at both timepoints (pooled acute injury studies vs. recovery studies).

From Table 3, there are no results that are statistically significant at the 0.05 level of significance (frontal p = 0.087, parietal p = 0.095 temporal = 0.494). Although the affected Parietal and Frontal lobe clearance rates do not overlap with normal controls, the differences do not reach statistical significance. The unaffected temporal lobe pooled data showed no evidence that the measures are different compared to control group clearance rates figure 2.

**Table 3.** Results of Linear Mixed Effects Models Comparing Acute mTBI Group Labeled Residual Proton Slopes to Controls.

| Lobe | Time | Mean | SE(M) | Difference | 95% CI (Diff) |  | t | df | p |
| --- | --- | --- | --- | --- | --- | --- | --- | --- | --- |
| Temporal | mTBI | -0.0525 | 0.0098 | 0.0109 | -0.0224 | 0.0442 | 0.70 | 14.1 | 0.494 |
|  | Control | -0.0634 | 0.0121 |  |  |  |  |  |  |
| Frontal | mTBI | -0.0455 | 0.0073 | 0.0215 | -0.0036 | 0.0465 | 1.85 | 13.0 | 0.087 |
|  | Control | -0.0670 | 0.0090 |  |  |  |  |  |  |
| Parietal | mTBI | -0.0416 | 0.0082 | 0.0230 | -0.0050 | 0.0520 | 1.80 | 12.9 | 0.095 |
|  | Control | -0.0651 | 0.0101 |  |  |  |  |  |  |

Figure 5 includes means and standard error bars of the slopes for subjects in each of the three groups for each lobe.

**Figure 5.**
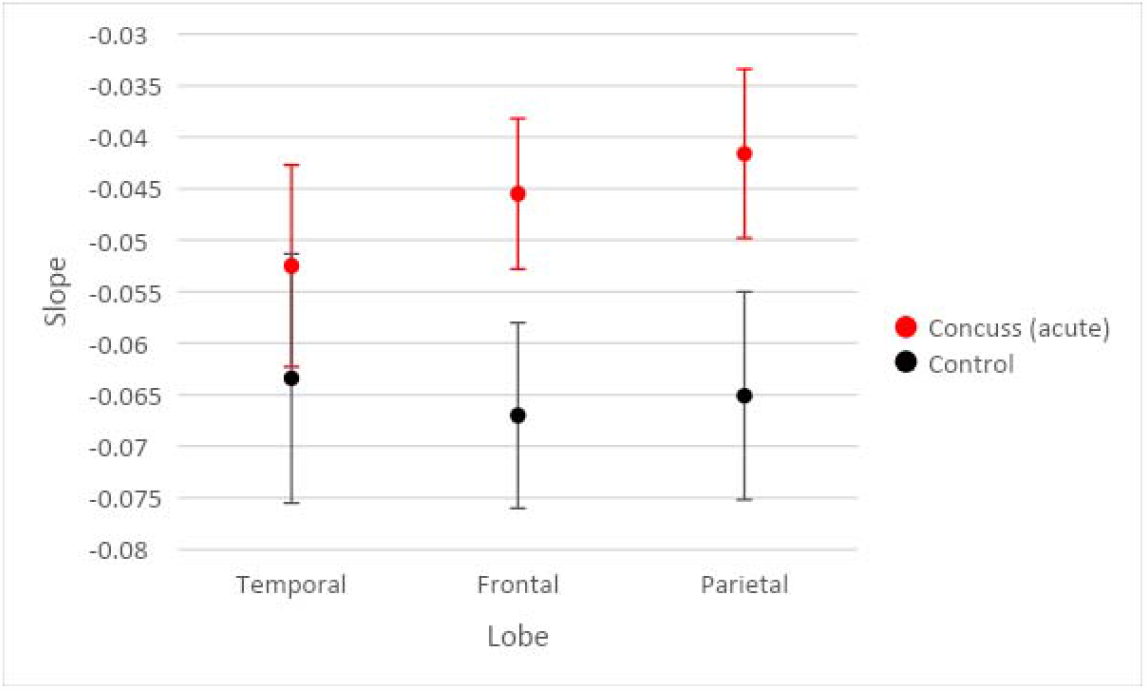
includes means and standard error bars of the GCR for subjects in each of the three groups for each lobe (pooled acute injury group vs control group)

Table 3 shows the results from the three linear mixed effects models comparing the slopes of the acute phase of the mTBI group to the control group.

From Table 4, there are no results that are statistically significant at the p≤ 0.05 level of significance. Thus, recovery clearance rates in the acute injury affected parietal, frontal, and temporal lobes showed no evidence that the measures are different from the control group [figure 3].

**Table 4.**
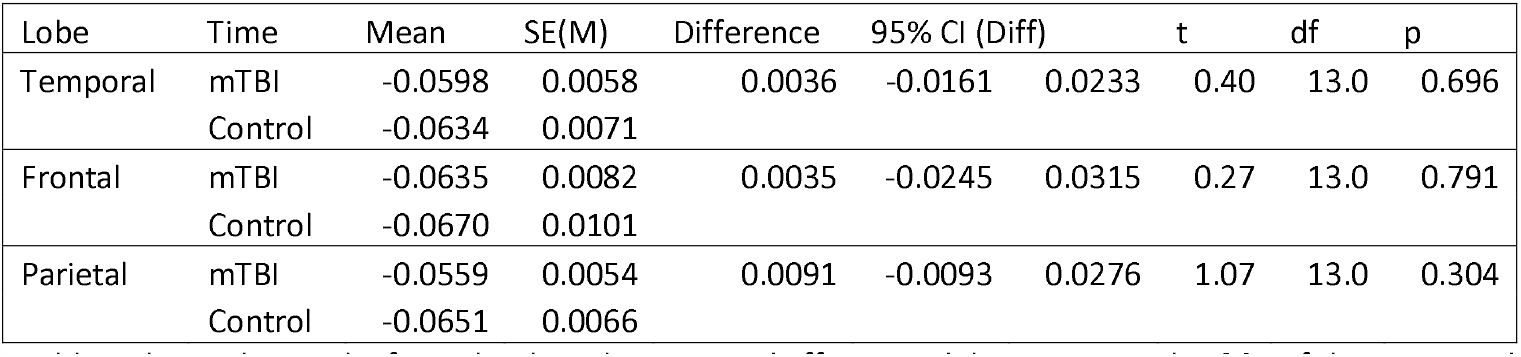
Results of Linear Mixed Effects Models Comparing Recovered mTBI Group Labeled Residual Proton Slopes to Control Group Table 4 shows the results from the three linear mixed effects models comparing the GCR of the recovered phase of the mTBI group to the control group.

Figure 6. Modeled Means and Standard Error Bars by Group and Lobe comparing GCR of recovery studies with control group studies.

**Figure 6.**
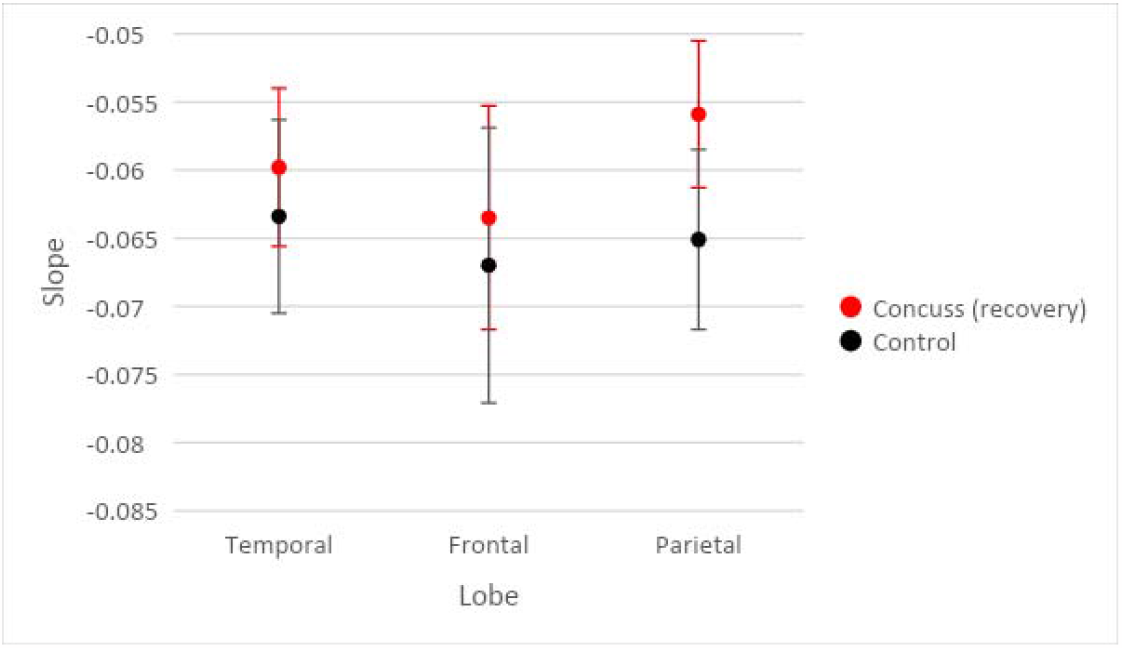
includes means and standard error bars of the GCR for subjects in each of the three groups for each lobe (pooled recovery studies vs. control studies).

Table 4 shows the results from the three linear mixed effects models comparing the slopes of the recovered phase of the mTBI group to the control group.

## Discussion

### Clinical Implications

This pilot study using ASL MRI within one week of acute mild head injury in student athletes with Glasgow Coma Scale score of 13-15 showed localized reduction in clearance rates (GCRs) of labeled protons^37^. When clinically cleared to return to their sport, statistically significant improvement of clearance rates were noted, suggesting good sensitivity of the technique. The GCRs improvement likely corresponds to repair of the BBB leak inherent to ACI. The advantages of this technique includes low cost, no exogenous contrast agents, MRI sequence availability, and short scan times. A larger study is warranted to further validate this approach for widespread use as a biomarker in ACI and recovery. With newer multi-labeling ASL techniques available, the scan time may be significantly reduced to only 5-6 minutes^38^. The advantage of this method is the use of intrinsic labeled protons in blood altered only by spin orientation, which does not disturb the underlying physiology. Gadolinium contrast techniques such as DCE require diffusion calculations and are invasive. Further, ASL MRI can be repeated multiple times given the absence of extrinsic contrast, which in contact sports activities, may provide guidance otherwise missing as to when and if an athlete can return safely to sport ^35,36^. Additionally, the technique may provide an outcome measure for various future therapeutic endeavors to repair BBB leak in more severe head injury and as well as a variety of diseases where it is impaired.

Of interest are the brain injury locations in this study showing the most common acute injury were the frontal and parietal lobes and with the exception of subject 5 sparing the temporal lobes. The mild degree and site of impact, likely explains this.

### Limitations

Limitations of our study include a small sample size (pilot study), and underrepresentation of male athlete volunteers. The latter is in part related to a higher risk of concussion in young women athletes related to several factors such as sex ^2, 26, 34^ hormones and physiologic changes. In addition, male athletes were less likely to volunteer. The small number of normal controls may explain the lack of statistical difference in the clearance rates compared with the acute injury results. Also, the mild nature of the head injuries may directly correlate with a less severe reduction in clearance rates. A larger study with inclusion of more serious head injury patients would be a logical next step. In absence of clearance rate recovery in more severe HI, anticipated cognitive neurologic deficits could be predicted, and appropriate rehab measures applied. Technical issues regarding low signal strength are compensated by averaging a large field of view in each of the brain regions studied and obtaining multiple time delay data points in the identical anatomic location.

This pilot study shows the potential of adopting 3D ASL MRI with further validation in the evaluation of ACI and presumed recovery as an available, noninvasive, inexpensive, and efficient reproducible biomarker.

## Data Availability

All data produced in the present study are available upon reasonable request to the authors
All data produced in the present work are contained in the manuscript.

## Acknowledgements

Charles Joseph, MD Lab

Liberty Athletics Team Physicians: Benjamin Ferry, MD, CAQSM, Mark Rolfs, DO, PharmD

Statistical analysis: Kim Love, PhD

MRI: Department of Radiology, Centra Lynchburg General Hospital, and Stan Gray, RPT.

## Data sharing

All result data is presented in the Appendix. Additional post analysis data is available upon request from the corresponding author.

## Contributorship

All authors contributed to the project execution and authorship of this manuscript.

## Patient project development involvement

Patients were not involved in the development of this project. Lay person involvement of this project was through the IRB committee approval process.

## Ethical approval

CENTRA IRB approval CHIRB0535

## Appendix

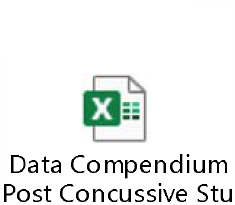

## Captions for figures 1-6

**Figure 1 caption** Composition of the ASL signal/time into its various components: arterial, capillary, and extra capillary spaces. TI (acquisition) times utilized in this study are between the red arrow limits. Note the signal composition is capillary and extra-capillary (interstitial) water, the latter is normally cleared by the glymphatic system The 7 PLDs or times of acquisition in his study are between the red arrow heads._______ total composite signal vs time -------- arterial signal contribution vs time -.-.-. intracapillary signal contribution vs time - - - - Extracapillary fluid signal contribution vs time.

T1 times (63% signal decay) of major signal contributors indicated by the colored dots.

Magenta dot = 800-850 msec T1 of white matter.

Orange dot = 1650 msec T1 of blood.

Green dot = 1700msec T1 of gray matter (all values are for 3T)

Dk blue dot = 3800msec T1 of water (CSF fluid); (all values are for 3T)

BAT = bolus arrival time; aaTT =artery-artery arrival time; τ = peak capillary arrival time

Adapted with publisher permission (Wiley) from reference 24.

Figure 2. Selected brain regions for ROI signal average. The blue oval contains the region of signal average: temporal lobe 650mm^2^; frontal lobe 1150mm^2^; and parietal lobe 760mm^2^. Homologous contralateral determinations were also obtained.

Figure 3 Example of signal averages vs. time. Acute phase temporal lobe and right Parietal lobe show GCR of zero or positive. The same regions upon recovery show negative GCR slopes within the normal range.

Figure 4. Modeled Means and Standard Error Bars by Time and Lobe.

Figure 5. Modeled Means and Standard Error Bars by Group and Lobe comparing recovery clearance rates with controls.

Figure 6. Modeled Means and Standard Error Bars by Group and Lobe comparing injury and control group clearance rates.

